# Early SARS-CoV-2 reinfections within 60 days highlight the need to consider antigenic variations together with duration of immunity in defining retesting policies

**DOI:** 10.1101/2022.04.04.22273172

**Authors:** Louis Nevejan, Lize Cuypers, Lies Laenen, Liselotte Van Loo, François Vermeulen, Elke Wollants, Ignace Van Hecke, Stefanie Desmet, Katrien Lagrou, Piet Maes, Emmanuel André

## Abstract

The emergence of the SARS-CoV-2 Omicron variant, characterized by a significant antigenic diversity compared to the previous Delta variant, had led to a decrease in antibody efficacy in both convalescent and vaccinees’ sera resulting in high number of reinfections and breakthrough cases worldwide. However, to date, reinfections are defined by the ECDC as two positive tests ≥60 days apart, influencing retesting policies after an initial positive test in several European countries. We illustrate by a clinical case supplemental by epidemiological data that early reinfections do occur within 60 days especially in young, unvaccinated individuals. In older patient groups, unvaccinated and patients with a basic vaccination scheme are more vulnerable to reinfections compared to patients who received a first booster vaccine. For this reason, we consider that the duration of protection offered by a previous infection should be reconsidered, in particular when a shift between consecutive SARS-CoV-2 variants occurs.

## Text

The sequential emergence of SARS-CoV-2 variants of concern (VOCs), characterized by both an antigenic drift and higher transmissibility, has been observed in all countries around the world at least three times during the last 13 months (1). These abrupt viral population replacements inevitably impact the residual population immunity, independently from the timing of a previous infection. While the SARS-CoV-2 Delta variant showed a limited antigenic diversity with previous VOCs, Omicron differs more significantly from other VOCs than any previous variant did at the time they emerged (2). The resulting decrease of antibody efficacy in both convalescent and vaccinees’ sera drives the high number of reinfection and breakthrough cases observed with Omicron compared to the observations made during the previous waves (3–6).

To date, reinfections with SARS-CoV-2 are defined by the European Centre for Disease Prevention and Control (ECDC) as two positive tests ≥60 days apart (technical report April 8, 2021) (7). This definition has influenced testing strategies in several countries, and many countries therefore do not recommend retesting patients for six months after a positive test. We consider that this definition needs to be revised.

To illustrate our point, we report here a case of an immunocompetent boy with no significant medical history, who was infected with SARS-CoV-2 Delta variant (≥7.0 log copies/mL, sublineage AY.43) in early December, 2021, concomitant with an outbreak at the patient’s school (Table 1). The patient’s sibling and one parent were infected as well (≥7.0 log copies/mL respectively ≥5.0 - <7.0 log copies/mL). All three developed mild COVID-19 symptoms. The other parent, boosted 1 month before, tested negative twice. Due to an unrelated trauma two weeks later, the patient was admitted to the hospital for surgery. Systematic SARS-CoV-2 screening performed at admission detected a low viral load (<3.0 log copies/mL), interpreted as viral remnant of the past infection 16 days earlier and typing of this sample was not possible due to low viral load. Three weeks later, the patient was readmitted for a second stage surgery. Preprocedural SARS-CoV-2 screening detected a strong positive result (5.1 log copies/mL) with Omicron BA.1 variant, therefore only 39 days after the patient’s infection with Delta. The patient remained pauci-symptomatic. High-risk contact screening of the patient’s brother detected a low viral load (<3.0 log copies/mL, untypable due to the low viral load), the mother tested SARS-CoV-2 negative and the father was not retested.

**Table 1.**
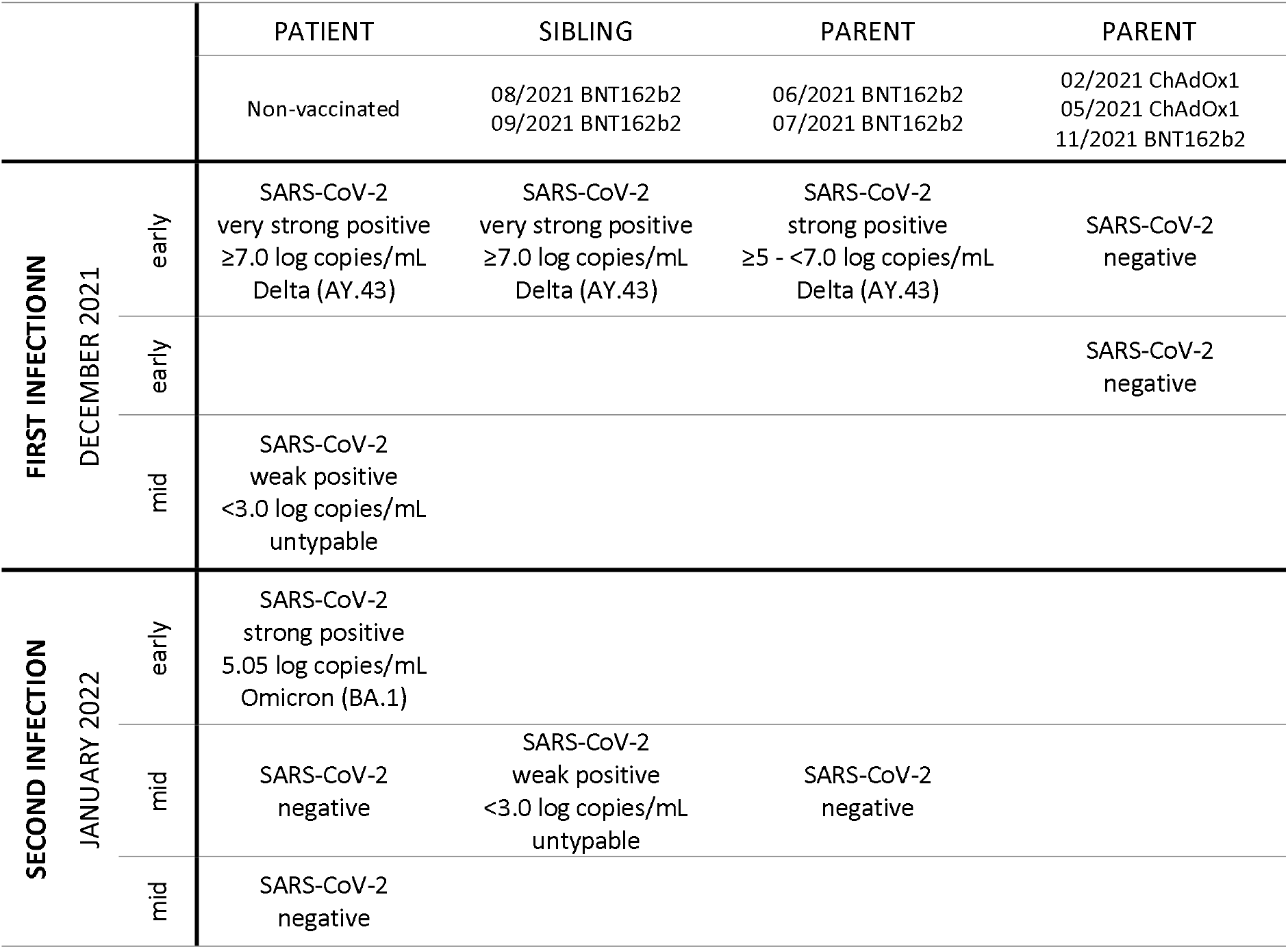
Results of SARS-CoV-2 screening and vaccination status in the patient and his family. All samples were analyzed using reverse transcription polymerase chain reaction, whole genome sequencing was performed on SARS-CoV-2 positive samples in the Belgian National Reference Center for Respiratory Pathogens (10).

Further, to put this clinical case into a wider epidemiological context, we estimated the incidence of early reinfections with 1) Omicron BA.1 after Delta infection and 2) with Omicron BA.2 after BA.1 infections in a community setting (Flemish Brabant, Belgium).

Between December 1, 2021 and February 7, 2022 (n = 9 weeks), a period characterized by the viral replacement of Delta by Omicron BA.1, a total of 56,831 ambulatory patients tested SARS-CoV-2 positive at the federal testing platform located in Leuven, Belgium. Among these, 0.16% (91 out of 56,831) had the S-gene detected in a first sample using the TaqPath PCR test (suggestive of Delta infection), while S-gene target failure (SGTF) was reported in a second positive sample within this time period (cycle threshold N gene <27.8), assuming a reinfection with Omicron BA.1 briefly after Delta infection.

Similarly, between January 1, 2022 and March 10, 2022 (n = 9 weeks), a period characterized by viral replacement of Omicron BA.1 by BA.2, a total of 48,829 patients tested SARS-CoV-2 positive. Among these, 0.01% (5 out of 48,829) presented an SGTF in a first sample but had an S-gene detected in a second positive sample. Considering the epidemiological context during the latter period in this region, reinfections with Omicron BA.2 after BA.1 infection were suspected in these patients.

In Figure 1, we report the age and vaccination status of these 96 patients with documented early reinfection, compared to the vaccination rate for the same age groups in the same geographic region (Flanders, Belgium) (8). Early reinfections were most frequently observed among young unvaccinated patients (<12 years old). In all age groups, patients with early reinfection had a lower vaccination rate compared to the corresponding age groups in the general population. Boosted patients had the lowest risk of early SARS-CoV-2 reinfection. Median time between the last vaccine dose and reinfection in all patients was 172 days (IQR 76-195 days). Median time between the two positive samples with different VOC was 47 days (range 17-65 days).

**Figure 1.**
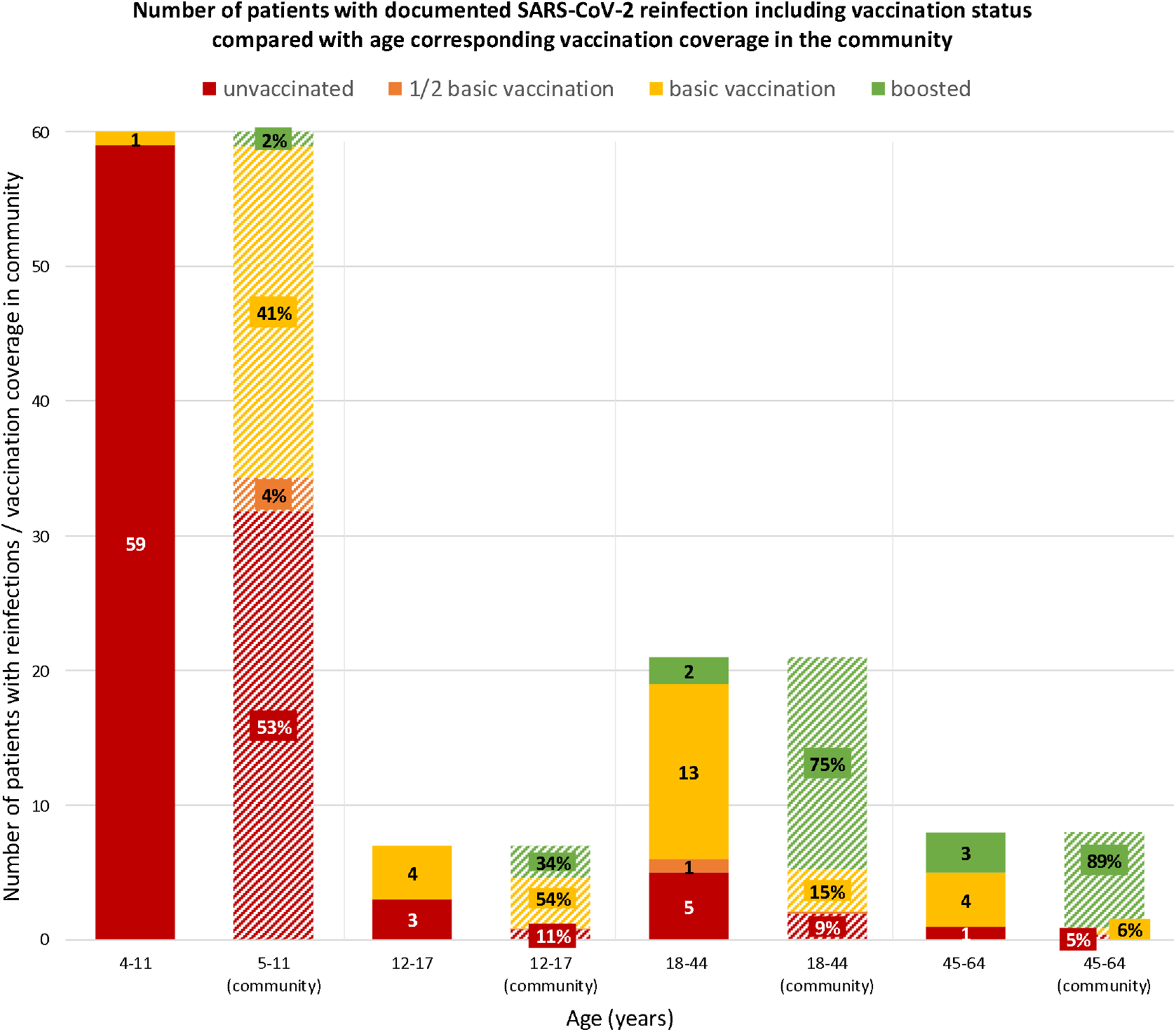
Number of patients with documented SARS-CoV-2 reinfection including vaccination status compared with age corresponding vaccination coverage in the community (i.e. Flanders, Belgium)

Previous large retrospective cohort studies showing a prolonged maintenance of protection against reinfection should be questioned in the context of Omicron (3). Our data confirms that early Omicron BA.1 reinfection (<60 days) after Delta infection and BA.2 reinfection after a BA.1 infection can occur, especially in young, unvaccinated individuals. In older patient groups, unvaccinated and patients with a basic vaccination scheme are more vulnerable to reinfections compared to patients who received a first booster vaccine. Data from Denmark (9) suggests reinfection usually results in mild disease not requiring hospitalization as is illustrated by this reported case.

For this reason, we consider that the duration of protection offered by a previous infection should be reconsidered, in particular when a shift between consecutive SARS-CoV-2 variants occurs. Testing policies should not assume that patients do not need to be retested within a period of 60 days, as reinfections, especially in unvaccinated young patients, are not inconceivable within this timeframe. It has been repeatedly observed that a full viral population replacement can occur in a matter of a few weeks, and such phenomenon will continue to impact the duration and efficacy of immunity in the future.

## Data Availability

All data produced in the present study are available upon reasonable request to the authors

## Acknowledgment

UZ Leuven, as national reference center for respiratory pathogens, is supported by Sciensano, which is gratefully acknowledged

## References

1. Mullen JL, Tsueng G, Abdel Latif A, Alkuzweny M, Cano M, Haag E, et al. Outbreak.info [Internet]. 2020 [cited 2022 Feb 4]. Available from: outbreak.info

2. Van der Straten K, Guerra D, Van Gils MJ, Bontjer I, Caniels TG, Van Willigen HDG, et al. Mapping the antigenic diversification of SARS-CoV-2. medRxiv [Internet]. 2022; Available from: https://www.medrxiv.org/content/10.1101/2022.01.03.21268582v1%0Ahttps://www.medrxiv.org/content/10.1101/2022.01.03.21268582v1.abstract

3. Kim P, Gordon SM, Sheehan MM, Rothberg MB. Duration of Severe Acute Respiratory Syndrome Coronavirus 2 Natural Immunity and Protection Against the Delta Variant: A Retrospective Cohort Study. Clin Infect Dis [Internet]. 2021 Dec 3;(Xx Xx):1–6. Available from: https://academic.oup.com/cid/advance-article/doi/10.1093/cid/ciab999/6448857

4. Planas D, Saunders N, Maes P, Guivel-Benhassine F, Planchais C, Buchrieser J, et al. Considerable escape of SARS-CoV-2 Omicron to antibody neutralization. Nature [Internet]. 2022 Feb 24;602(7898):671–5. Available from: https://www.nature.com/articles/s41586-021-04389-z

5. Pulliam JRC, Schalkwyk C van, Govender N, Gottberg A von, Cohen C, Groome MJ, et al. Increased risk of SARS-CoV-2 reinfection associated with emergence of the Omicron variant in South Africa. medRxiv [Internet]. 2021; Available from: https://www.medrxiv.org/content/10.1101/2021.11.11.21266068v2%0Ahttps://www.medrxiv.org/content/10.1101/2021.11.11.21266068v2.abstract

6. Rahman S, Rahman MM, Miah M, Begum MN, Sarmin M, Mahfuz M, et al. COVID-19 reinfections among naturally infected and vaccinated individuals. Sci Rep [Internet]. 2022 Dec 26;12(1):1438. Available from: https://www.nature.com/articles/s41598-022-05325-5

7. European Centre for Disease Prevention and Control. Reinfection with SARS-CoV-2: implementation of a surveillance case definition within the EU/EEA [Internet]. Stockholm; 2021. Available from: https://www.ecdc.europa.eu/en/publications-data/reinfection-sars-cov-2-implementation-surveillance-case-definition-within-eueea

8. Vaesen J. Covid Vaccinaties België [Internet]. 2022 [cited 2022 Mar 21]. Available from: https://covid-vaccinatie.be/nl

9. Stegger M, Edslev SM, Sieber RN, Ingham AnC, Ng KL, Tang M-HE, et al. Occurrence and significance of Omicron BA. 1 infection followed by Corresponding author. medRxiv [Internet]. 2022; Available from: https://doi.org/10.1101/2022.02.19.22271112

10. Pirnay J-P, Selhorst P, Hong SL, Cochez C, Potter B, Maes P, et al. Variant Analysis of SARS-CoV-2 Genomes from Belgian Military Personnel Engaged in Overseas Missions and Operations. Viruses [Internet]. 2021 Jul 13;13(7):1359. Available from: https://www.mdpi.com/1999-4915/13/7/1359

